# Impact of COVID-19 in Individuals with Autism Spectrum Disorders: Analysis of a National Private Claims Insurance Database

**DOI:** 10.1101/2021.03.31.21254434

**Authors:** Arun Karpur, Vijay Vasudevan, Andy Shih, Thomas Frazier

**Author notes:** Correspondence regarding this paper should be directed to Arun Karpur at. No external funding was received for this research. Author note: AK conceptualized the study, supervised data analysis, wrote the manuscript, TF conceptualized and contributed to the writing, VV contributed to the writing of manuscript, and AS helped with conceptualization of the study. Authors also acknowledge contributions from FAIR Health Inc., specifically Ali Russo, Eric Okurowski, David Cheng, and Josh Smolinsky. Arun Karpur is the Director, Data Science and Evaluation Research, Autism Speaks at Princeton NJ office.

## Abstract

The COVID-19 pandemic continues to have a detrimental impact on individuals with disabilities. Data from FAIR Health’s FH® NPIC (National Private Insurance Claims) database,^1^ one of the nation’s largest databases of private insurance claim records, were analyzed to understand the experiences of individuals with ASD in COVID-19 pandemic. Multivariate logistic regression models revealed that individuals with ASD + ID were nine times more likely to be hospitalized (OR = 9.3; 95% CI: 6.9 – 12.5) and were nearly six times more likely have an elevated length of hospital stay(OR = 5.9; 95% CI: 3.5 – 10.1) compared those without ASD + ID. These findings point to their need for prioritization in access to vaccines for preventing COVID-19 infection and morbidities.

---

The global pandemic from SARS-CoV-2 virus infection causing the Coronavirus disease in 2019 (COVID-19) has led to wide ranging crises in health and social supports for vulnerable individuals, especially for those with developmental disabilities and autism spectrum disorders (ASD). A study of individuals with ASD in Germany illustrated that the COVID-19 pandemic increased ASD-related behavioral difficulties, reduced sleep quality, and increased hypersensitivity (Mutluer, Doenyas, & Aslan Genc, 2020). People with ASD also had difficulty understanding of the need for social distancing and staying at home, consistently using personal protective equipment (e.g., masks, gloves, etc.). Further, their caregivers did not have access to resources to educate their child with ASD about COVID-19 and ways to prevent infections. Finally, the study illustrated that increases in ASD-related behavioral challenges were correlated with severity of anxiety in parents and caregivers. Similarly, another European study illustrated an exacerbation of pre-existing behavioral challenges in individuals with ASD, as well as parent reported challenges in managing free time and structured activities, during the COVID-19 pandemic (Colizzi et al., 2020). Children with ASD that did not receive services from schools experienced more intense behavioral problems.

Beyond the direct impact on behavior and physical health of individuals with ASD, the COVID-19 pandemic surfaced several related challenges, such as food insecurity and mental health problems among households (Jeste et al., 2020). As educational and public health institutions grapple with optimal protocols for school openings in the U.S., data on clinical experiences including prevalence of COVID-19 and hospitalization during the COVID-19 pandemic for individuals with ASD are weak to nonexistent. It is postulated that since individuals with ASD have higher concentration of proinflammatory cytokines(Jyonouchi, 2013; Jyonouchi, Geng, Rose, Bennuri, & Frye, 2019), seen in other chronic conditions that predispose individuals to COVID-19, the prevalence of COVID-19 infection in likely higher. The overlap in neuroinflammatory pathophysiology in ASD and COVID-19, could possibly exacerbate challenging behaviors and mental health problems (Lima, Barros, & Aragao, 2020). A recent case series of sixteen patients with ASD with COVID-19 disease indicated that while most hospitalized individuals with ASD exhibited common symptoms of the infection, several had idiosyncratic manifestations contributing to challenges in diagnosis and treatment (Nollace et al., 2020).

Turk and colleagues, in their analysis utilizing the TriNetX COVID-19 Research Network Data, consisting of electronic medical records from 42 health systems in the U.S., illustrated that the case fatality rates for individuals with intellectual and developmental disabilities (IDD) was comparable to the general population. However, they also identified an age-related trend where the case fatality was higher among younger individuals with IDD than those without IDD, and it is likely due to higher proportion of comorbidities among young individuals with individuals with IDD (Turk, Landes, Formica, & Goss, 2020). While it was not reported, it is likely that the study sample included many individuals with ASD grouped under the broad definition of IDD, as nearly 40% of individuals with ASD have cooccurring ID (Maenner, Shaw, Baio, & al., 2020). To our knowledge, no prior study has examined the experiences of hospitalization, including the duration of hospital stay among individuals with ASD. Thus, the purpose of this analysis is to illustrate the impact of COVID-19 infection on health of individuals with ASD when compared to their peers with other chronic conditions.

## Methods

### Data source

Claims records from February 1, 2020 through September 30, 2020 from the FAIR Health National Private Insurance Claims (FH NPIC^©^) database were utilized for this analysis. The Centers for Medicare and Medicaid Services (CMS) Chronic Conditions Data Warehouse (CCW) definitions were applied to the diagnostic codes attached to the claims data. Additionally, diagnostic codes for each claim were examined retrospectively until October 1, 2017 to allow for capturing diagnosis of ASD and other listed CCW conditions from prior visits (for more information see: https://www2.ccwdata.org/web/guest/condition-categories). Professional and facility claims were examined to understand the prevalence of COVID-19 infection. A cohort of 35,898,076 patients who incurred a claim between February 1, 2020 and September 30, 2020 were analyzed. The following condition groups were created based on the diagnostic codes as dummy variables (1 = has this condition; 0 = does not have this condition):

*Autism Spectrum Disorders (ASD only); Autism Spectrum Disorders AND Intellectual Disabilities and Related Conditions (ASD + ID); Autism Spectrum Disorders AND Other Developmental Delays (ASD + DD); Intellectual Disabilities and Related Conditions; Attention-deficit/Hyperactivity disorder(ADHD), Conduct Disorders, and Hyperkinetic Syndrome; Learning Disabilities; Other Developmental Delays; Hypertension; Chronic Kidney Disease; Hyperlipidemia; Anemia; Diabetes; Ischemic Heart Disease; Heart Failure; Obesity; Rheumatoid Arthritis/Osteoarthritis; Chronic Obstructive Pulmonary Disease; Atrial Fibrillation; Depressive Disorders; Anxiety Disorders; None of the Specified Conditions*.

Specifically, claims with birth-related disorders (e.g., ASD, ID, DD) were assigned to their respective groups based on birth-related disorders to create mutually exclusive claim records. Claims that were likely to be adult-onset chronic conditions were grouped into their respective diagnostic category without being mutually exclusive as it was difficult to ascertain their primary diagnosis given the length of clinical history claims available for the analysis.

### Statistical analyses

Any claim with a COVID-19 diagnosis resulting from hospitalization or a visit to doctor’s office was recorded as a case of COVID-19. Descriptive analysis studied distribution in trends in hospitalization in the reported claims. Elevated length of stay was a categorical variable - longer than median length of stay for all claims and shorter than median length of stay. Two separate stepwise multiple logistic regression models were created to study the likelihood of hospitalization and experiencing elevated length of hospital stay. Each of the condition groups (e.g., ASD + ID, ASD + DD, ASD, etc.), coded as dummy variables, were entered along with gender (male; female) and age-groups (0-18 years; 19 – 29 years; 30 – 39 years; 40 – 49 years; 50 – 59 years; 60 – 69 years; 70 years and older) using the step-wise forward logistic regression with a significance level of 0.3 required to allow the variable into the model and a significance level of 0.35 required to allow the variable to stay into the model with each subsequent entry.

## Results

Based on the analytical dataset, the overall prevalence of COVID-19 infection was estimated to be 2.04%. Out of the total 144,147 individuals with ASD identified in the FAIR Health database who incurred a claim between February 2020 and September 2020, 1,300 individuals had a positive COVID-19 diagnosis resulting in an overall prevalence of 0.90%. A greater proportion of individuals with ASD + ID, ASD + DD, and ASD only were males and belonged to young age-groups (i.e., 0 – 18 years, and 19 – 29 years) compared with other condition groups. The estimated prevalence of COVID-19 infection was 2.01% among individuals with ASD + ID (201 cases out of 9,991 individuals), 0.75% among individuals with ASD + DD (538 cases out of 71,999 individuals), and 0.90% among individuals with ASD only (561 cases out of 62,165 individuals). The highest prevalence of COVID-19 was among individuals with heart failure without ASD or other developmental conditions (4.30%). The overall mortality from COVID-19 for individuals in the analytical data set was 0.89%. The proportion of deaths or mortality from COVID-19 in infected individuals was 2.49% among individuals with ASD + ID, 0.37% among individuals with ASD + DD, and 0.89% for individuals with ASD only. Individuals with heart failure without ASD or other developmental conditions had the highest proportion of mortality from COVID-19 (10.5%). Table 1 illustrates the proportion of prevalence and mortality for the condition groups. The unadjusted data (Figure 1) indicate larger proportions of outpatient doctor’s office visits for individuals with ASD + ID, ASD + DD, and ASD only (69.2%, 80.3%, and 85.9% respectively). Further, as the proportion utilizing doctor’s office declined, the proportion of claims with hospitalization increased.

**Table 1.** Prevalence of COVID-19 infection and mortality in condition groups

| **Condition Groups** | **Prevalence Proportion** | **Mortality Proportion** |
| --- | --- | --- |
| Autism Spectrum Disorders AND Other Developmental Delays | 0.75% | 0.37% |
| Learning Disabilities | 0.85% | 1.21% |
| Autism Spectrum Disorders | 0.90% | 0.89% |
| Other Developmental Delays | 0.95% | 0.57% |
| None of the Specified Conditions | 1.57% | 0.02% |
| ADHD, Conduct Disorders, and Hyperkinetic Syndrome | 1.94% | 0.48% |
| Autism Spectrum Disorders AND Intellectual Disabilities and Related Conditions | 2.01% | 2.49% |
| Intellectual Disabilities and Related Conditions | 2.21% | 5.44% |
| Depressive Disorders | 2.51% | 2.02% |
| Anxiety Disorders | 2.70% | 1.45% |
| Rheumatoid Arthritis/ Osteoarthritis | 2.76% | 2.45% |
| Hyperlipidemia | 2.92% | 2.00% |
| Hypertension | 2.98% | 2.77% |
| Atrial Fibrillation | 3.19% | 8.87% |
| Ischemic Heart Disease | 3.35% | 6.49% |
| Obesity | 3.51% | 1.85% |
| Diabetes | 3.60% | 3.19% |
| Chronic Obstructive Pulmonary Disease | 3.74% | 5.23% |
| Chronic Kidney Disease | 3.81% | 7.17% |
| Anemia | 4.11% | 3.47% |
| Heart Failure | 4.30% | 10.50% |

Note: Data table is sorted in the descending order of the prevalence proportion.

**Figure 1.**
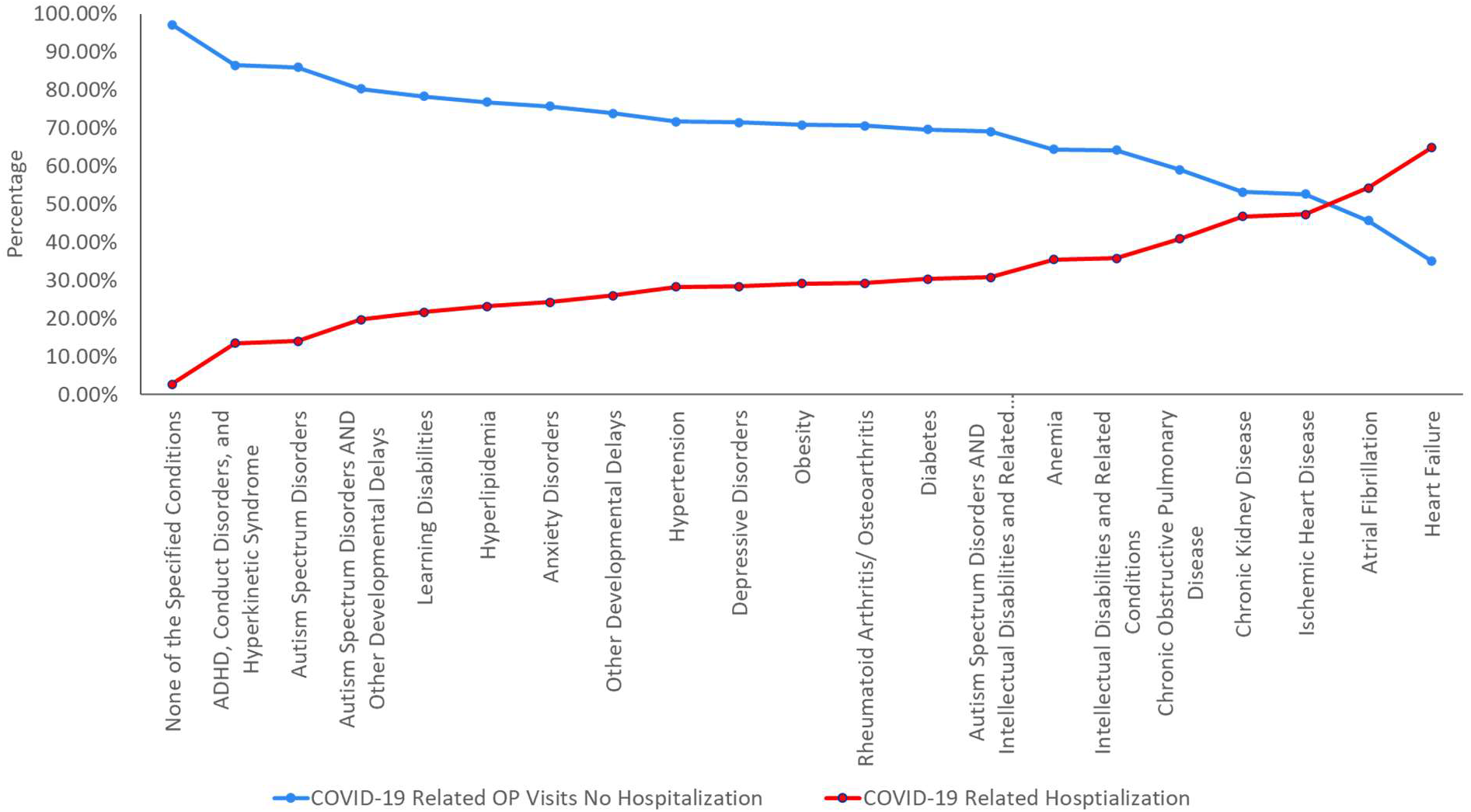
Frequency of outpatient visits and hospitalizations in COVID-19 for condition groups

The overall proportion of hospitalization for individuals with COVID-19 infection in the analytical data set was 12.70%. Figure 2 illustrates the odds ratios for hospitalization for each of the condition groups based on the stepwise forward logistic regression model. After adjusting for age group and gender, individuals with ASD + ID were more than nine times likely to be hospitalized compared with those that did not have this condition (OR = 9.3; 95% CI: 6.9 – 12.5). Similarly, individuals with ASD + DD were nearly six times more likely (OR = 5.9; 95% CI: 4.7 – 7.3) and those with ASD only were nearly four times more likely to be hospitalized (OR = 3.6; 95% CI: 2.9 – 4.6) compared to those that did not have this condition. The overall proportion of individuals with elevated length of of hospital stay (i.e., longer than median length) from COVID-19 infection was 5.58%. Figure 3 illustrates the odds ratios for elevated length of hospital stay for each of the condition groups based on the stepwise forward logistic regression model. Individuals with ASD + ID were six times more likely to have elevated length of hospital stay compared to those that did not have this condition (OR = 5.9; 95% CI: 3.5 – 10.1). Individuals with ASD + DD were four times more likely (OR = 4.0; 95% CI: 2.7 – 5.9), and those with ASD only were nearly three times more likely to have elevated length of hospital stay (OR = 2.8; 95% CI: 1.8 – 4.5) compared to those that did not have this condition.

**Figure 2.**
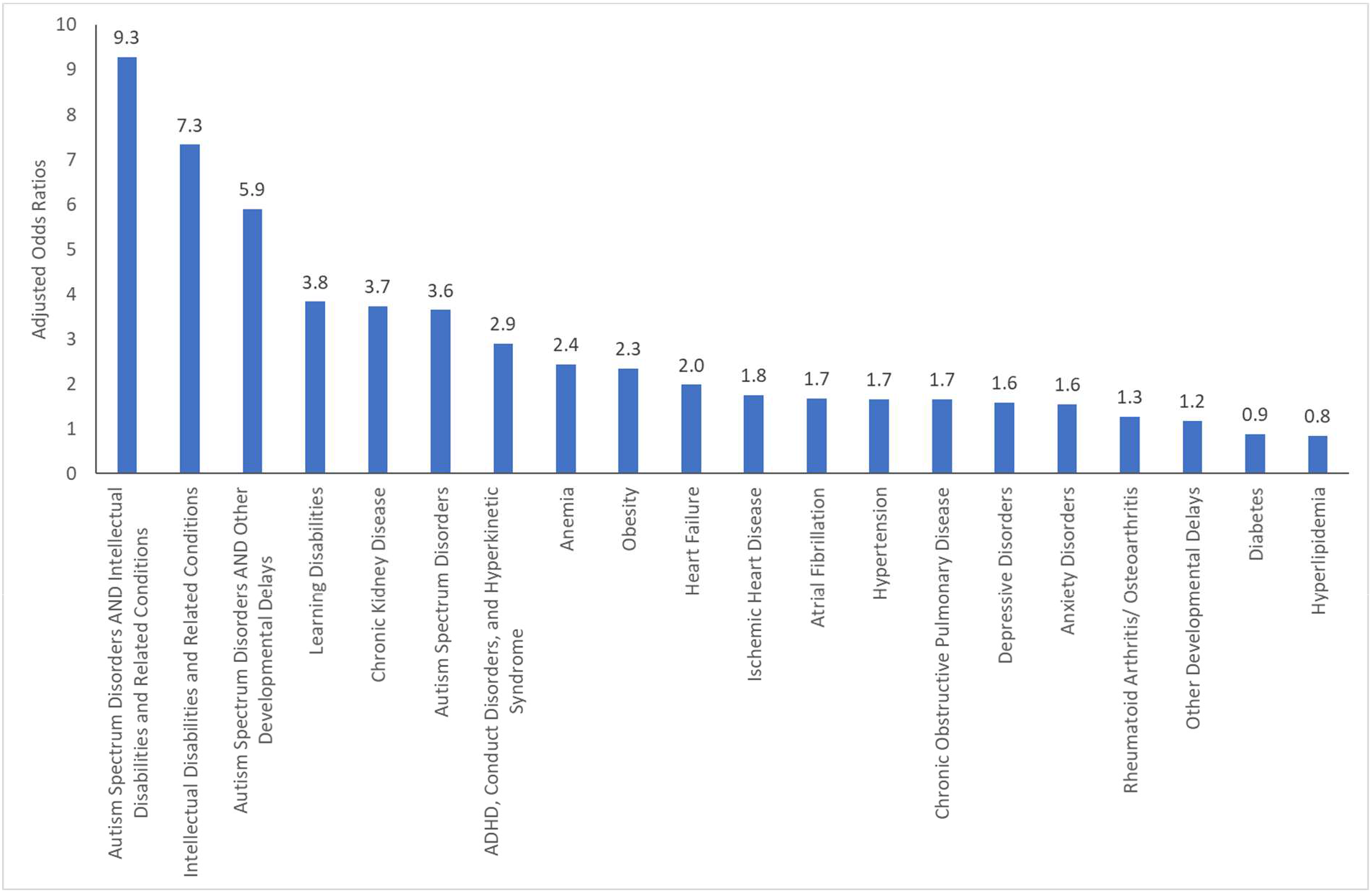
Likelihood of hospitalization in COVID-19 infection across the condition groups

**Figure 3.**
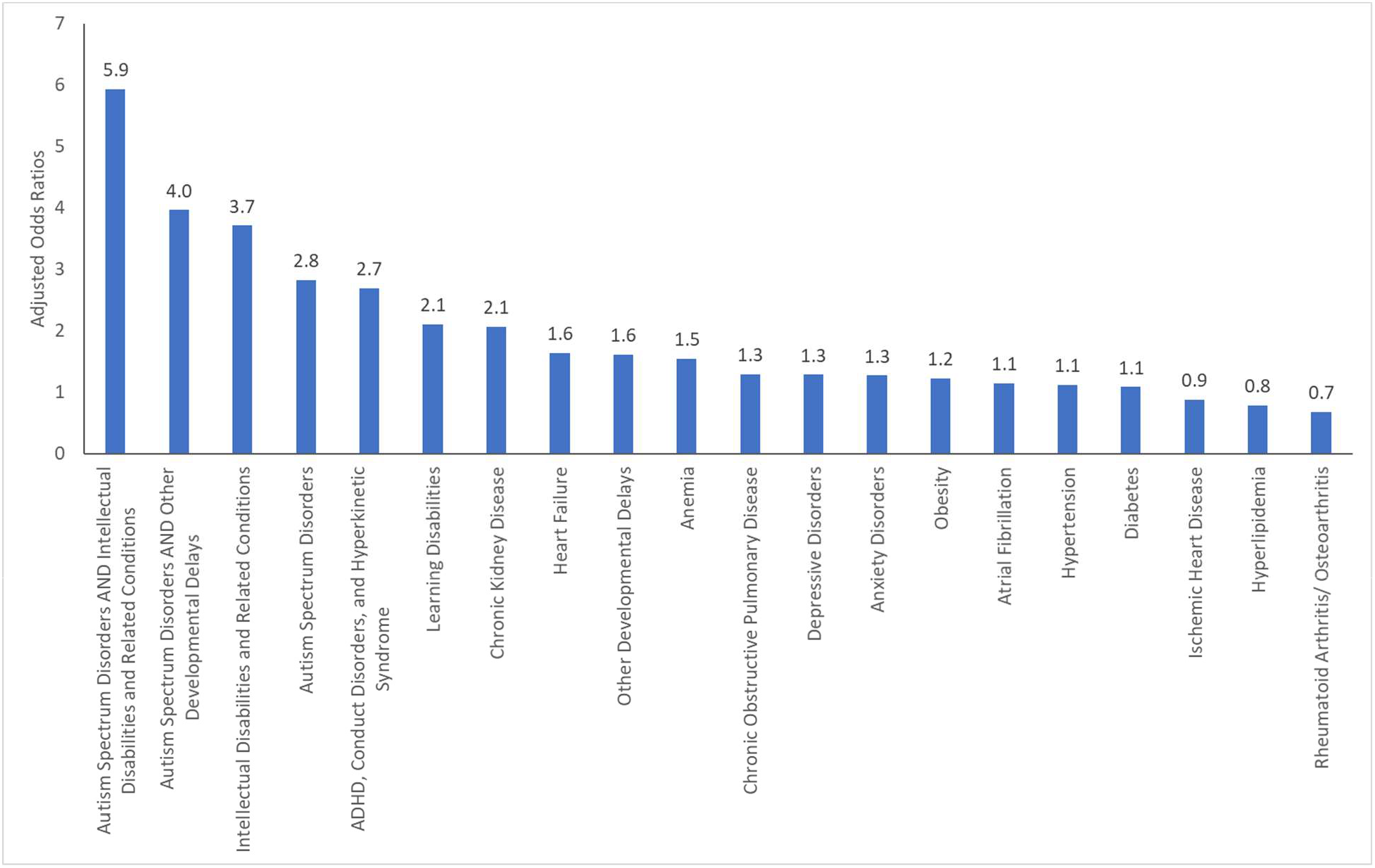
Likelihood of higher than median length of stay in hospital from COVID-19 infection

## Discussion

This is a first study to illustrate a higher likelihood of hospitalization and elevated length of hospital stay in COVID-19 for individuals with ASD and other comorbidities. While a lower proportion of individuals with ASD acquired COVID-19 infection compared to other condition groups, they were significantly more likely to be hospitalized for COVID-19 and have longer length of stay in the hospital. Contrastingly, individuals with other chronic conditions had higher prevalence of COVID-19 infections (e.g., Heart Failure, Obesity, Diabetes) had lower adjusted odds of hospitalization and elevated length of hospital stay. A lower overall prevalence of COVID-19 in ASD is likely driven by underlying demographic patterns. A closer examination of the demographic distribution indicates that more than 90% of individuals with ASD were in the younger age range (i.e., less than 30 years old), of which nearly 70% belonged to 0 – 18-year-old group. As a result, most individuals with ASD in the data are likely living with their families during pandemic and as a result are less likely to acquire COVID-19 infection than individuals in other settings such as congregate living environments. A substantial amount of reporting of higher morbidity and mortality in COVID-19 among individuals with disabilities, especially for those with intellectual and developmental disabilities have been associated with their living situation in congregate care settings (Landes, Turk, Formica, McDonald, & Stevens, 2020).

Once individuals with ASD acquire COVID-19 infection, the regression models indicate that they have a higher likelihood of hospitalization and elevated length hospital stay. While Cunningham and colleagues (2020) described a similar pattern of lower COVID-19 prevalence and higher morbidity among young individuals using electronic medical records data, the quantitative differences in the likelihood of hospitalization and duration of stay for individuals with ASD is concerning and requires attention (Cunningham et al., 2020). It is known that individuals with ASD, in general, have higher medical needs resulting from cooccurring mental health and other conditions such as epilepsy, digestive disorders, etc. (Karpur, Lello, Frazier, Dixon, & Shih, 2018; Shea et al., 2018) It is possible that the COVID-19 infection, irrespective of severity, precipitates behavior health challenges leading to hospitalization and longer duration of stay (Righi et al., 2017). Further, Several immunological theories, including the more recent preposition of abnormal melatonin production among individuals ASD, might contributed to the increased severity of COVID-19 infection (Brown et al., 2021).

The findings of increased morbidity in COVID-19 are relevant from the perspective of increasing discussion on prioritization of populations for COVID-19 vaccines. Only a handful of states have considered prioritizing individuals with intellectual disabilities (see: https://www.kff.org/policy-watch/the-next-phase-of-vaccine-distribution-high-risk-medical-conditions/) and this does not include all individuals with ASD. However, given the challenges in consistently implementing social distancing practices, and use of PPEs among individuals with ASD it would be useful to also include them as one of the high-risk populations for immunizations.

These analyses are limited by a few factors. Firstly, the FH NPIC^©^ data are limited to just private health claims data, while many people with ASD are on public insurance like Medicaid. Medicaid funds both healthcare and long-term services and supports such as congregate or community-based housing for people with ASD or other developmental disabilities. Future studies should explore hospitalization and length of stay because of COVID-19 for people with ASD who have Medicaid. Secondly, the findings are based on claims data that trigger a record upon receipt of services and the record for this study extends only to September 2020. It is possible that the prevalence of COVID-19 infection is much higher than is reported here in the analysis and it certainly increased substantially, as infections increased in late fall/early winter. An additional limitation is that condition identification was based on the ICD-10 diagnostic codes that may miss some individuals. Further, a substantial proportion of private claims data do not report on race/ethnicity, making it difficult to examine disparities in experiences (Ng, Ye, Ward, Haffer, & Scholle, 2017). Factors indicative of social and economic vulnerability, including living situation, would likely present more nuanced and policy-relevant findings. Future studies that employ natural language processing or medical record review to identify COVID-19, people with ASD, and other important factors related to hospitalization and length of stay would be useful for extending the present work and further characterizing the impact of COVID-19 on the ASD population.

## Data Availability

Research for this article is based upon healthcare claims data compiled and maintained by FAIR Health, Inc. The authors are solely responsible for the conclusions reflected in this article. FAIR Health, Inc. is not responsible for the any of the opinions expressed in this article.

## Footnotes

1. Research for this article is based upon healthcare claims data compiled and maintained by FAIR Health, Inc. The authors are solely responsible for the conclusions reflected in this article. FAIR Health, Inc. is not responsible for the any of the opinions expressed in this article.

## Supplementary Materials

**Table 1.** Results from stepwise forward logistic regression model for hospitalization in COVID-19

| **Demographics and Clinical Characteristics** | **Level** | **Estimate** | **Odds Ratio** | **95% Wald Confidence Interval** | **P value** |
| --- | --- | --- | --- | --- | --- |
| **Age Band** | 0 to 18 years | -0.3255 | 0.563 | 0.541-0.586 | <.0001 |
|  | 19 to 29 years | -0.2155 | 0.628 | 0.611-0.646 | <.0001 |
|  | 30 to 39 years | Ref |  |  | <.0001 |
|  | 40 to 49 years | -0.1631 | 0.662 | 0.645-0.680 | <.0001 |
|  | 50 to 59 years | -0.0241 | 0.761 | 0.742-0.781 | 0.0043 |
|  | 60 to 69 years | 0.168 | 0.922 | 0.896-0.949 | <.0001 |
|  | 70 and Over | 0.311 | 1.064 | 1.024-1.105 | <.0001 |
| **Gender** | Female | Ref |  |  |  |
|  | Male | -0.0142 | 0.972 | 0.957-0.988 | 0.0005 |
| **Only Autism Spectrum Disorders** | Absent | Ref |  |  |  |
|  | Present | 0.6469 | 3.647 | 2.871-4.632 | <.0001 |
| **Autism with Intellectual Disabilities and Related Conditions** | Absent | Ref |  |  |  |
|  | Present | 1.114 | 9.281 | 6.867-12.544 | <.0001 |
| **Autism with other Development Disorders** | Absent | Ref |  |  |  |
|  | Present | 0.8877 | 5.902 | 4.764-7.313 | <.0001 |
| **ADHD, Conduct Disorders, and Hyperkinetic Syndrome** | Absent | Ref |  |  |  |
|  | Present | 0.5313 | 2.894 | 2.765-3.029 | <.0001 |
| **Intellectual Disabilities and Related Conditions** | Absent | Ref |  |  |  |
|  | Present | 0.9959 | 7.329 | 6.371-8.431 | <.0001 |
| **Learning Disabilities** | Absent | Ref |  |  |  |
|  | Present | 0.6716 | 3.831 | 3.349-4.382 | <.0001 |
| **Other Developmental Delays** | Absent | Ref |  |  |  |
|  | Present | 0.0838 | 1.182 | 0.950-1.471 | 0.1328 |
| **Hypertension** | Absent | Ref |  |  |  |
|  | Present | 0.2535 | 1.66 | 1.627-1.695 | <.0001 |
| **Chronic Kidney Disease** | Absent | Ref |  |  |  |
|  | Present | 0.6585 | 3.732 | 3.647-3.820 | <.0001 |
| **Hyperlipidemia** | Absent | Ref |  |  |  |
|  | Present | -0.0874 | 0.84 | 0.823-0.857 | <.0001 |
| **Anemia** | Absent | Ref |  |  |  |
|  | Present | 0.4453 | 2.436 | 2.389-2.485 | <.0001 |
| **Diabetes** | Absent | Ref |  |  |  |
|  | Present | -0.0676 | 0.874 | 0.853-0.894 | <.0001 |
| **Ischemic Heart Disease** | Absent | Ref |  |  |  |
|  | Present | 0.2812 | 1.755 | 1.704-1.807 | <.0001 |
| **Heart Failure** | Absent | Ref |  |  |  |
|  | Present | 0.3416 | 1.98 | 1.900-2.064 | <.0001 |
| **Obesity** | Absent | Ref |  |  |  |
|  | Present | 0.4244 | 2.337 | 2.295-2.380 | <.0001 |
| **Rheumatoid Arthritis / Osteoarthritis** | Absent | Ref |  |  |  |
|  | Present | 0.1179 | 1.266 | 1.238-1.294 | <.0001 |
| **Chronic Obstructive Pulmonary Disease and Bronchitis** | Absent | Ref |  |  |  |
|  | Present | 0.252 | 1.655 | 1.603-1.709 | <.0001 |
| **Atrial Fibrillation** | Absent | Ref |  |  |  |
|  | Present | 0.26 | 1.682 | 1.614-1.753 | <.0001 |
| **Depression** | Absent | Ref |  |  |  |
|  | Present | 0.2279 | 1.577 | 1.539-1.617 | <.0001 |
| **Anxiety Disorders** | Absent | Ref |  |  |  |
|  | Present | 0.2204 | 1.554 | 1.520-1.589 | <.0001 |

Note: Data source FAIR Health National Private Insurance Claims (FH NPIC®). C-statistic = 0.805.

**Table 2.** Results from stepwise forward logistic regression model for mean length of stay in hospital in COVID-19

| **Demographics and Clinical Characteristics** | **Level** | **Estimate** | **Odds Ratio** | **95% Wald Confidence Interval** | **P value** |
| --- | --- | --- | --- | --- | --- |
| **Age Band** | 0 to 18 years | -0.28 | 1.365 | 1.252-1.488 | <.0001 |
|  | 19 to 29 years | -0.5395 | 1.053 | 0.988-1.122 | <.0001 |
|  | 30 to 39 years | Ref |  |  |  |
|  | 40 to 49 years | 0.0126 | 1.829 | 1.734-1.929 | 0.4751 |
|  | 50 to 59 years | 0.3106 | 2.464 | 2.341-2.593 | <.0001 |
|  | 60 to 69 years | 0.4787 | 2.915 | 2.758-3.081 | <.0001 |
|  | 70 and Over | 0.6089 | 3.32 | 3.113-3.541 | <.0001 |
| **Gender** | Female | Ref |  |  |  |
|  | Male | 0.2521 | 1.656 | 1.608-1.705 | <.0001 |
| **Only Autism Spectrum Disorders** | Absent | Ref |  |  |  |
|  | Present | 0.5202 | 2.83 | 1.799-4.451 | <.0001 |
| **Autism with Intellectual Disabilities and Related Conditions** | Absent | Ref |  |  |  |
|  | Present | 0.8904 | 5.935 | 3.499-10.067 | <.0001 |
| **Autism with other Development Disorders** | Absent | Ref |  |  |  |
|  | Present | 0.6889 | 3.966 | 2.684-5.861 | <.0001 |
| **ADHD, Conduct Disorders, and Hyperkinetic Syndrome** | Absent | Ref |  |  |  |
|  | Present | 0.4944 | 2.688 | 2.463-2.933 | <.0001 |
| **Intellectual Disabilities and Related Conditions** | Absent | Ref |  |  |  |
|  | Present | 0.6556 | 3.711 | 2.942-4.680 | <.0001 |
| **Learning Disabilities** | Absent | Ref |  |  |  |
|  | Present | 0.3713 | 2.102 | 1.673-2.640 | <.0001 |
| **Other Developmental Delays** | Absent | Ref |  |  |  |
|  | Present | 0.2371 | 1.607 | 1.119-2.307 | 0.0102 |
| **Hypertension** | Absent | Ref |  |  |  |
|  | Present | 0.0547 | 1.116 | 1.075-1.157 | <.0001 |
| **Chronic Kidney Disease** | Absent | Ref |  |  |  |
|  | Present | 0.3632 | 2.068 | 1.997-2.141 | <.0001 |
| **Hyperlipidemia** | Absent | Ref |  |  |  |
|  | Present | -0.1252 | 0.779 | 0.752-0.806 | <.0001 |
| **Anemia** | Absent | Ref |  |  |  |
|  | Present | 0.2186 | 1.549 | 1.501-1.598 | <.0001 |
| **Diabetes** | Absent | Ref |  |  |  |
|  | Present | 0.0404 | 1.084 | 1.046-1.124 | <.0001 |
| **Ischemic Heart Disease** | Absent | Ref |  |  |  |
|  | Present | -0.0625 | 0.883 | 0.846-0.921 | <.0001 |
| **Heart Failure** | Absent | Ref |  |  |  |
|  | Present | 0.2452 | 1.633 | 1.551-1.720 | <.0001 |
| **Obesity** | Absent | Ref |  |  |  |
|  | Present | 0.0994 | 1.22 | 1.183-1.258 | <.0001 |
| **Rheumatoid Arthritis / Osteoarthritis** | Absent | Ref |  |  |  |
|  | Present | -0.1953 | 0.677 | 0.653-0.702 | <.0001 |
| **Chronic Obstructive Pulmonary Disease and Bronchitis** | Absent | Ref |  |  |  |
|  | Present | 0.1293 | 1.295 | 1.237-1.355 | <.0001 |
| **Atrial Fibrillation** | Absent | Ref |  |  |  |
|  | Present | 0.0696 | 1.149 | 1.090-1.212 | <.0001 |
| **Depression** | Absent | Ref |  |  |  |
|  | Present | 0.1255 | 1.285 | 1.235-1.338 | <.0001 |
| **Anxiety Disorders** | Absent | Ref |  |  |  |
|  | Present | 0.1198 | 1.271 | 1.224-1.319 | <.0001 |

Note: Data source FAIR Health National Private Insurance Claims (FH NPIC®). C-statistic = 0.724.

